# Antipsychotics effects on network-level reconfiguration of cortical morphometry in first-episode schizophrenia

**DOI:** 10.1101/2021.01.17.21249965

**Authors:** Yuchao Jiang, Yingchan Wang, Huan Huang, Hui He, Yingying Tang, Wenjun Su, Lihua Xu, Yanyan Wei, Tianhong Zhang, Hao Hu, Jinhong Wang, Jijun Wang, Cheng Luo, Dezhong Yao

**Author notes:** Corresponding to: University of Electronic Science and Technology of China, Second North Jianshe Road, Chengdu, 610054, China; (C. Luo). These two authors contributed equally to this work (Yuchao Jiang, Yingchan Wang). These authors contributed equally to this work (Jijun Wang, Cheng Luo).

## Abstract

**Background:** Cortical thickness reductions are evident in patients with schizophrenia. Associations between antipsychotic medications (APMs) and cortical morphometry have been explored in schizophrenia patients. This raises the question of whether the reconfiguration of morphological architecture by APM plays potential compensatory roles for abnormalities in the cerebral cortex.

**Methods:** Structural MRI were obtained from 127 medication-naive first-episode schizophrenia (FES) patients and 133 matched healthy controls. Patients received 12 weeks of APM and were categorized as responders (n=75) or nonresponders (n=52) at follow-up. Using surface-based morphometry and structural covariance analysis, this study investigated the short-term effects of antipsychotics on cortical thickness and cortico-cortical connectivity. Global efficiency was computed to characterize network integration of the large-scale structural connectome. The relationship between connectivity and cortical thinning was examined by the structural covariance analysis among top-n regions with thickness reduction.

**Results:** Widespread cortical thickness reductions were observed in pre-APM patients. Post-APM patients showed more reductions in cortical thickness, even in the frontotemporal regions without baseline reductions. Covariance analysis revealed strong cortico-cortical connectivity and higher network integration in responders than in nonresponders. Notably, the nonresponders lacked key nodes of the prefrontal and temporal regions for the covariance network between top-n regions with cortical thickness reductions.

**Conclusions:** Antipsychotic effects are not restricted to a single brain region but rather exhibit a network-level covariance pattern. Neuroimaging connectomics highlights the positive effects of antipsychotics on the reconfiguration of brain architecture, suggesting that abnormalities in regional morphology may be compensated by increasing interregional covariance when symptoms are controlled by antipsychotics.

## Introduction

Schizophrenia (SZ) has been hypothesized to be a neurodevelopmental disorder that is typically characterized onset in late adolescence or early adulthood ^1^. Structural magnetic resonance imaging (MRI) studies suggest widespread brain structural abnormalities even in the earliest stages of SZ ^2^. Investigations of brain structure have revealed progressive reductions in gray matter volume with longer illness duration, beginning in the thalamus and progressing to the frontal regions and then to the temporal and occipital lobes ^3^. Additionally, coordinated gray matter loss exhibits a pattern of irregular topographic distribution across cerebral cortices in SZ ^4^. This evidence supports the link between brain morphological changes involved in neuropathological processes and systematic ‘dysconnectivity’ (involving insufficient or ineffective communications) at the network level in SZ ^5^.

Structural covariance network (SCN) analysis is frequently employed to assess the network-level disturbances in structural connections across brain regions in SZ ^6^. Cortico-cortical connectivity, established by structural covariance (SC), reflects interindividual differences in brain structures (i.e., cortical thickness reductions) and covaries between distant cortical regions across the population. SC has been associated with some kind of brain connections, such as the anatomical connectivity of white matter fiber tractography or the functional connectivity of synchronous neuronal activation ^6, 7^. A plausible mechanism to interpret SC is that synapses between distant neurons can have a mutually trophic or protective effect on neuronal development, leading to strong covariance in their morphology ^6^. It is suggestive of the ‘common fate’ of brain regions in terms of cortical reorganization processes when brain plastic changes occur during antipsychotic medications ^5^.

In SZ, SC analysis has been used to identify alterations across cortices in frontal, temporal and parietal regions ^4^. Frontal-temporal brain areas also exhibit cortical thinning in SZ, possibly due to overpruning of synapses during adolescence ^8^. Importantly, the strength of frontal–temporal covariance has been associated with auditory hallucinations ^9^, which is one of the most common symptoms in SZ. Graph theoretical studies also reported reduced integrity of the SCN in SZ ^10^, suggesting a low network efficiency of brain topological organization. Thus, it is understandable that these distributed changes in cortical thickness are indicative of a plastic reorganization process occurring in response to antipsychotics.

The association of antipsychotic medications (APM) with cortical morphometry has been explored in SZ studies. A meta-analysis of longitudinal studies involving 1,155 SZ patients and 911 healthy subjects showed higher loss of cortical total volume, which was related to cumulative antipsychotic intake ^11^. In addition to the reduction in total volume, findings associated with APM effects focused on frontal, temporal and parietal lobes ^12^. In addition, altered volumes in subcortical regions were associated with symptomatic improvement following APM treatment ^13^. Although these studies have identified APM effects on certain brain regions, they ignored the existence of coordinated relationships between brain regions. Interregional covariance reflects developmentally mediated dysconnectivity and plays a critical role in the pathophysiologic trajectory of brain abnormalities in SZ ^5, 6^. More recent work has used morphometric covariance analysis to examine brain topological reorganization in medicated patients with SZ ^14^. They found that nonresponders to treatment exhibited worse network integration than responders ^14^. This raises the question of whether the reconfiguration of the structural architecture by APM-induced morphological plasticity plays potential compensatory functions for abnormalities in the cerebral cortex, which is primed for relief of clinical symptoms in SZ. This hypothesis should be investigated in a large cohort of individuals with medication-naive first-episode SZ (FES) who have high homogeneity in illness duration, symptom severity and treatment approaches. In addition, a longitudinal design study is needed to clarify the short-term effect of APM on brain morphometry.

To address it, we estimated the region-wise cortical thickness and large-scale SCN in 127 medication-naive FES patients and 133 healthy subjects. Patients were grouped on the basis of their response to APM at follow-up. The aim of this study was to explore (1) whether the APM effects are not constrained within a single brain region but exhibit a topographical distribution across brain regions and (2) whether cortico-cortical connectivity inferred from the SCN could explain the extent and location of cortical thickness reductions following APM. We hypothesized that APM responders would show enhanced integration in the SCN and stronger cortico-cortical connectivity among regions with cortical thickness reductions compared with nonresponders.

## Materials and Methods

### Participants

A total of 127 patients with medication-naive FES (SZ group) who were diagnosed using the Diagnostic and Statistical Manual of Mental Disorders, 4th Edition (DSM-IV) and 133 healthy controls (HC group) matched for age, gender, education and handedness were recruited from Shanghai Mental Health Center. The exclusion criteria are described in the Supplementary Materials. MRI was scanned at baseline for all subjects and at follow-up after 12 weeks of APM for only patients. Following the baseline scanning, patients received second-generation antipsychotics (Supplementary Materials). At follow-up, patients were grouped as responders (SR group, n=75) and nonresponders (NR group, n=52) by the criterion of less than 50% the Positive and Negative Syndrome Scale (PANSS) reduction for nonresponse ^16^. The study was approved by the Institutional Review Board of Shanghai Mental Health Center. All subjects/patients signed written informed consent forms.

### Image data acquisition

High-spatial-resolution T1-weighted structural images were collected by a 3-Tesla MRI (Siemens MR B17). Acquisition parameters are provided in the Supplementary Materials.

### Image processing and cortical thickness estimation

To characterize the surface-based morphometry of the cerebral cortex, structural images were processed with the Computational Anatomy Toolbox within SPM12 (http://www.neuro.uni-jena.de/cat/). Briefly, a fully automated processing procedure included tissue segmentation and estimation of cortical thickness and central surface based on the projection-based thickness (PBT) method ^17^. Next, topology correction ^18^, spherical mapping and registration ^19^ was performed. The cortical surface was then parcellated into 360 regions of interest (ROIs) based on the HCP Multi-Modal Parcellation ^20^. Each ROI’s properties are documented in Supplementary Table S1. Finally, ROI-wise cortical thickness was extracted from individual native space using the CAT12’s ROI tools.

### Statistical analysis

#### Demographic differences

Demographic and clinical information differences were tested using t-tests for the continuous variables between the SZ and HC groups and between the two patient groups. The chi-square test was used for the categorical variables.

#### Cortical thickness differences

At baseline, we compared the baseline cortical thickness between all SZ patients and HCs using a two-sample t-test. ANOVA and post hoc t-tests were performed to compare the differences among the SR, NR and HC groups. To examine cortical thickness changes after APM, two-way repeated measures ANOVA and post hoc paired t-test were conducted in the SR and NR groups. The univariate statistical test was implemented independently for each of 360 cortical regions while controlling for age, square of age, gender, years of education and total intracranial volume (TIV) ^4^. To correct for multiple comparisons and to minimize the bias of data distribution, a permutation-based procedure was considered for controlling the family wise error (FWE) rate ^21^ (Supplementary Materials).

#### Associations with treatment outcomes

To examine the relationship between cortical thickness changes and treatment outcomes, the differences between the follow-up and baseline were extracted from each region that exhibited significant cortical thickness reduction at the follow-up. A linear stepwise regression analysis was performed to assess the association between these regional reductions and PANSS total score reductions in each patient group.

### Structural covariance network

SC analysis has been a robust and widely used method to characterize interregional structural connectivity between cortical distant regions ^6^, based on the assumption that intersubject differences in cortical morphology covary across individuals between distant regions that are anatomically connected^22^.

SCN was constructed using the cortical thickness change. Briefly, for each patient, the difference in cortical thickness between baseline and follow-up was computed as cortical thickness change. Cortical thickness change of each ROI was further corrected by eliminating the bias of gender, education, age, and the square of age, as well as TIV by a regression model ^6^. Pearson’s correlation coefficients were calculated across the subjects in each group between pairwise ROIs and was further transformed into a z value to improve normality. This yielded a correlation matrix that quantified the SC between all pairs of ROIs for each patient group. To reduce the number of correlations of false positive, false discovery rate correction was performed (P<0.000001).

To further investigate the network integration of the SCN, global efficiency was used for the measurement of network topology. The global efficiency of a network is denoted by the averaged inverse shortest path length and has been regarded as a superior measure of integration ^23^. The null hypothesis of equality in the global efficiency of the SCN between the SR and NR groups was tested using a permutation test (Supplementary Materials).

### Structural covariance in top-n regions with cortical thickness reduction

To investigate the impact of varying severity of cortical thickness change at follow-up, we analyzed the SC in top-n regions with cortical thickness change (as there was no thickness increase at the follow-up compared with the baseline, the ‘thickness change’ refers in particular to thickness reduction after the treatment). The methodology has been reported in a prior study ^4^ and is described briefly here. First, regions were ranked from highest to lowest according to the severity of cortical thickness changes at the follow-up, which was assessed by Cohen’s d effect size. Subsequently, the averaged SC among the top-n regions with the highest cortical thickness changes (i.e., SC_top-n_) was calculated using the thickness change values. The mean SC was also computed in a set of randomly chosen n regions (i.e., SC_rand-n_). Randomly chosen sets were repeated 5,000 and thus generated a distribution of SC of random n regions. According to the location of the actual SC_top-n_ within the distribution of SC_rand-n_ values, a p value was obtained for the null hypothesis of equality in mean SC among top-n regions and random regions. This analysis was repeated independently for n=2 to the maximum threshold (i.e., 360) to explore the impact of varying severity of cortical thickness changes (i.e., a larger n included regions with smaller cortical thickness reductions). As n increased, continuous values of mean SC among top-n regions were computed and could be plotted as a function of n. The area under the curve (AUC) of this plot was used to calculate a global p value for all n values. To determine which the top n exhibited the strongest SC compared to the randomly chosen n regions, the SC_top-n_ was transformed into a z score by subtracting the average of SC_rand-n_ values and then dividing it by the standard deviation of SC_rand-n_ values.

### Reproducibility and ancillary analysis

Reproducibility and ancillary analysis are provided in the Supplementary Materials.

## Results

### Demographic characteristics

Demographic information is described in Table 1.

**Table 1.** Demographic and clinical data of participants.

| (A) | Schizophrenia | Healthy controls | P value |
| --- | --- | --- | --- |
| Subject Number | 127 | 133 |  |
| Age (years) | 24.61(7.0) | 23.7(5.9) | 0.254<sup>a</sup> |
| Gender (Male/Female) | 63/64 | 66/67 | 0.998<sup>b</sup> |
| Education (years) | 12.9(2.9) | 13.5(2.8) | 0.110<sup>a</sup> |
| Handness (left/right) | - | - |  |
| TIV (mm<sup>3</sup>) | 1480.5(148.8) | 1476.9(133.0) | 0.838<sup>a</sup> |
| Illness duration (months) | 13.4(16.0) | - | - |
| CPZ (mg/d) | 402.8(188.9) | - | - |
| Baseline PANSS |  |  |  |
| Positive score | 24.0(5.1) | - | - |
| Negative score | 19.1(6.7) | - | - |
| General score | 42.4(6.9) | - | - |
| Total score | 85.8(12.9) | - | - |
| 12-weeks PANSS |  |  |  |
| Positive score | 12.6(4.1) | - | - |
| Negative score | 14.4(4.8) | - | - |
| General score | 28.8(5.7) | - | - |
| Total score | 55.8(12.5) | - | - |
| PANSS reduction (%) | 53.0(21.1) | - | - |
| (B) | Responds | Nonresponds | P value |
| Subject Number | 75 | 52 |  |
| Age (years) | 25.3(6.6) | 23.6(7.4) | 0.176<sup>a</sup> |
| Gender (Male/Female) | 35/40 | 28/24 | 0.426<sup>b</sup> |
| Education (years) | 13.4(2.9) | 12.3(2.7) | 0.033<sup>a</sup> |
| Handness (left/right) | - | - |  |
| TIV (mm<sup>3</sup>) | 1478.1(147.6) | 1483.9(152.0) | 0.832<sup>a</sup> |
| Illness duration (months) | 13.2(15.0) | 13.7(17.4) | 0.871<sup>a</sup> |
| CPZ (mg/d) | 406.2(197.9) | 397.9(176.9) | 0.810<sup>a</sup> |
| Baseline PANSS |  |  |  |
| Positive score | 24.6(5.0) | 23.2(5.3) | 0.125<sup>a</sup> |
| Negative score | 18.4(6.6) | 20.2(6.7) | 0.146<sup>a</sup> |
| General score | 43.6(7.2) | 40.8(6.0) | 0.020<sup>a</sup> |
| Total score | 86.9(14.1) | 84.1(10.9) | 0.221<sup>a</sup> |
| 12-weeks PANSS |  |  |  |
| Positive score | 11.0(3.0) | 14.9(4.3) | <0.001<sup>a</sup> |
| Negative score | 12.0(3.6) | 17.8(4.3) | <0.001<sup>a</sup> |
| General score | 26.0(4.6) | 32.8(4.9) | <0.001<sup>a</sup> |
| Total score | 49.0(9.2) | 65.7(9.6) | <0.001<sup>a</sup> |
| PANSS reduction (%) | 66.7(12.8) | 33.3(13.7) | <0.001<sup>a</sup> |

<sup>a</sup> P values were obtained by using two sample t-tests.
<sup>b</sup> P values were obtained by using the chi-square test.
TIV, Total Intracranial Volume; CPZ, Chlopromazine equivalents; PANSS, Positive And Negative Syndrome Scale.

### Cortical thickness baseline comparisons

Compared with the HCs, a total of 73, 90 and 12 regions were observed to present significantly thinner cortical thickness in the baseline SZ (combined SRt1 and NRt1), SR and NR groups, respectively (FWE-corrected P<0.05). Increased cortical thickness was not found in the three patient groups. In the SR group, thinner cortical thickness illustrated a distributed pattern involving frontal, parietal, temporal and occipital lobes as well as posterior cingulate cortex (Figure 1). In the NR group, thinner cortical thickness was observed in local regions of the frontal and parietal cortex (Figure 1). Details of regions with significant cortical thickness reductions are described in Supplementary Tables S2-S4.

**Figure 1.**
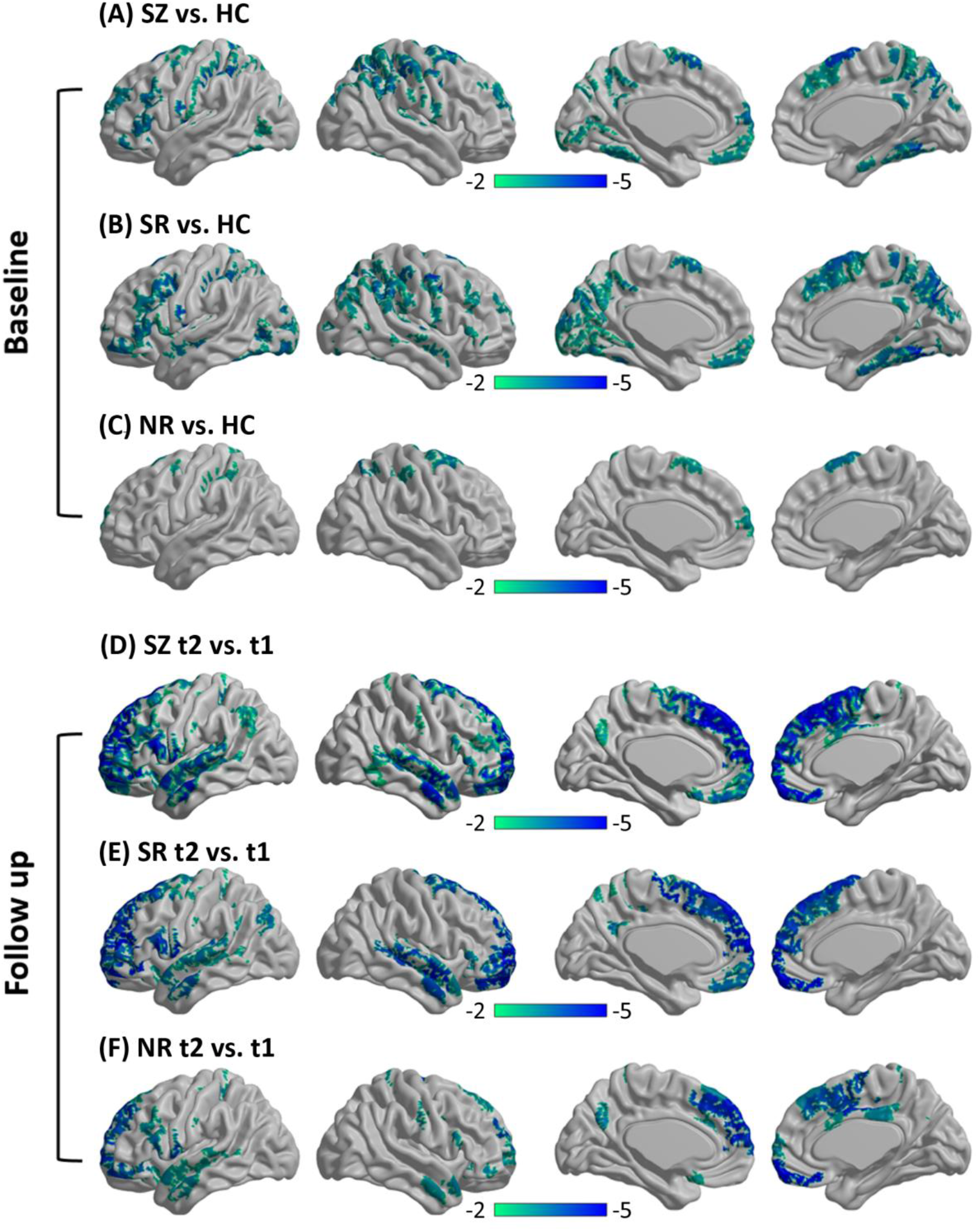
Cortical thickness comparisons. At the baseline, thinner cortical thickness are shown in the three patient groups (A, all SZ patients; B, SR patients; C, NR patients) compared with HC. At the follow-up (t2), cortical thickness reductions are shown in the two patient groups (D, all SZ patents; E, SR patients; F, NR patients) compared with the baseline (t1). Regions for which the null hypothesis was rejected after controlling the FWE rate at 5% are shown in color maps. Color bar represents the t-values.

### Cortical thickness changes

Compared with the baseline, the SR group at follow-up exhibited more extensive cortical thickness changes (only reductions) in the medial frontal cortex and right temporal regions, although cortical thickness changes were found in extensive regions (all SZ patients group, n=85; SR group, n=73; NR group, n=43) for both patient groups (FWE-corrected P<0.05) (Figure 1). Regions with significant cortical thickness changes at the follow-up and effect size are listed in Supplementary Tables S5-S7. The interaction effect between group and time was not evident after multiple comparisons correction.

### Associations between cortical thickness changes and treatment outcomes

Regression analysis found a significant relationship between the cortical thickness change in the left area-45 of the inferior frontal cortex and PANSS total reduction (T=2.37, P<0.05), which was only observed in the SR group (Figure S1). In addition, a supplementary analysis (Supplementary Materials) indicated that the relationship was not affected by gender, age, education, TIV, CPZ, illness duration or baseline PANSS total scores.

### Structural covariance network

Figure 2A and 2B show surviving SC connections after multiple comparisons correction (FDR-corrected P<0.000001) for the SR and NR groups, respectively. A larger number of surviving connections was observed in the SR group than in the NR group by the permutation test (P=0.020). In the network of the SR group, the majority of these connections (63.04%) were associated with frontal and parietal cortices, as shown in Figure S2. In addition, the null hypothesis of equality in network integration between the SR and NR groups was rejected, indicating superior global efficiency in the SR group by the permutation test (P=0.029) (Figure 3C).

**Figure 2.**
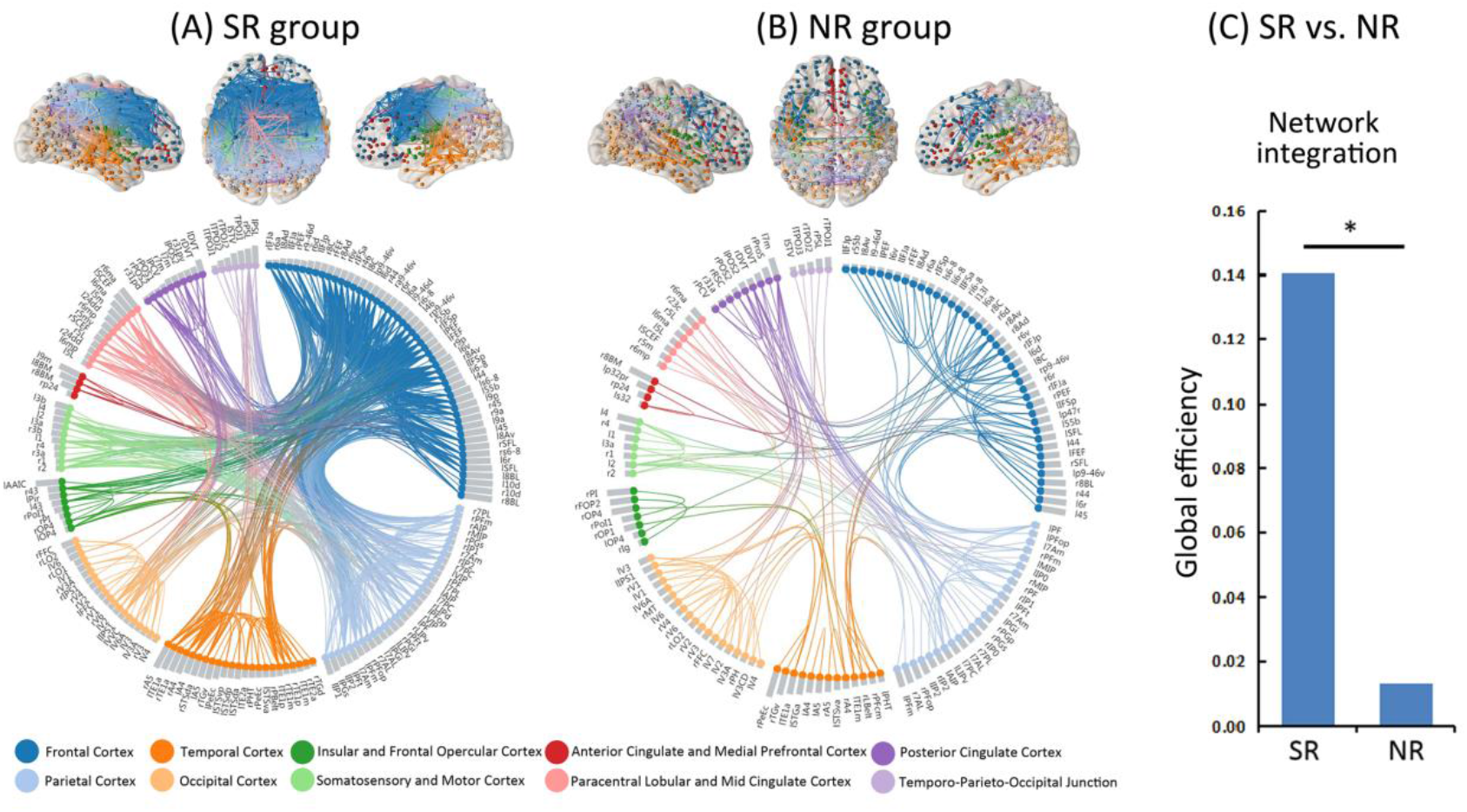
Cortico-cortical connectivity of structural covariance network is shown by brain connectivity maps^a^ and a circular connectogram^b^ for the SR group (panel A) and NR group (panel B), after multiple comparisons correction by the false discovery rate corrected P<0.000001. (C) The null hypothesis of equality in network integration between the SR and NR groups was rejected by the permutation test (P<0.05), indicating superior global efficiency in the SR group compared with the NR group. **Note:** ^a^ Brain regions are colored on the brain connectivity maps according to cortex classifications (described below the circular connectogram). Left hemisphere is shown on the left side of the axial map. ^b^ Brain regions are grouped on the circular connectogram according to cortex classifications (described below the circular connectogram). Within each sub-group, regions are ranked according to the severity of cortical thickness reductions at the follow-up, assessed by Cohen’d effect size (i.e., grey column adjacent to nodes). Brain connectivity maps and circular connectogram were produced by using NeuroMArVL (http://immersive.erc.monash.edu.au/neuromarvl).

**Figure 3.**
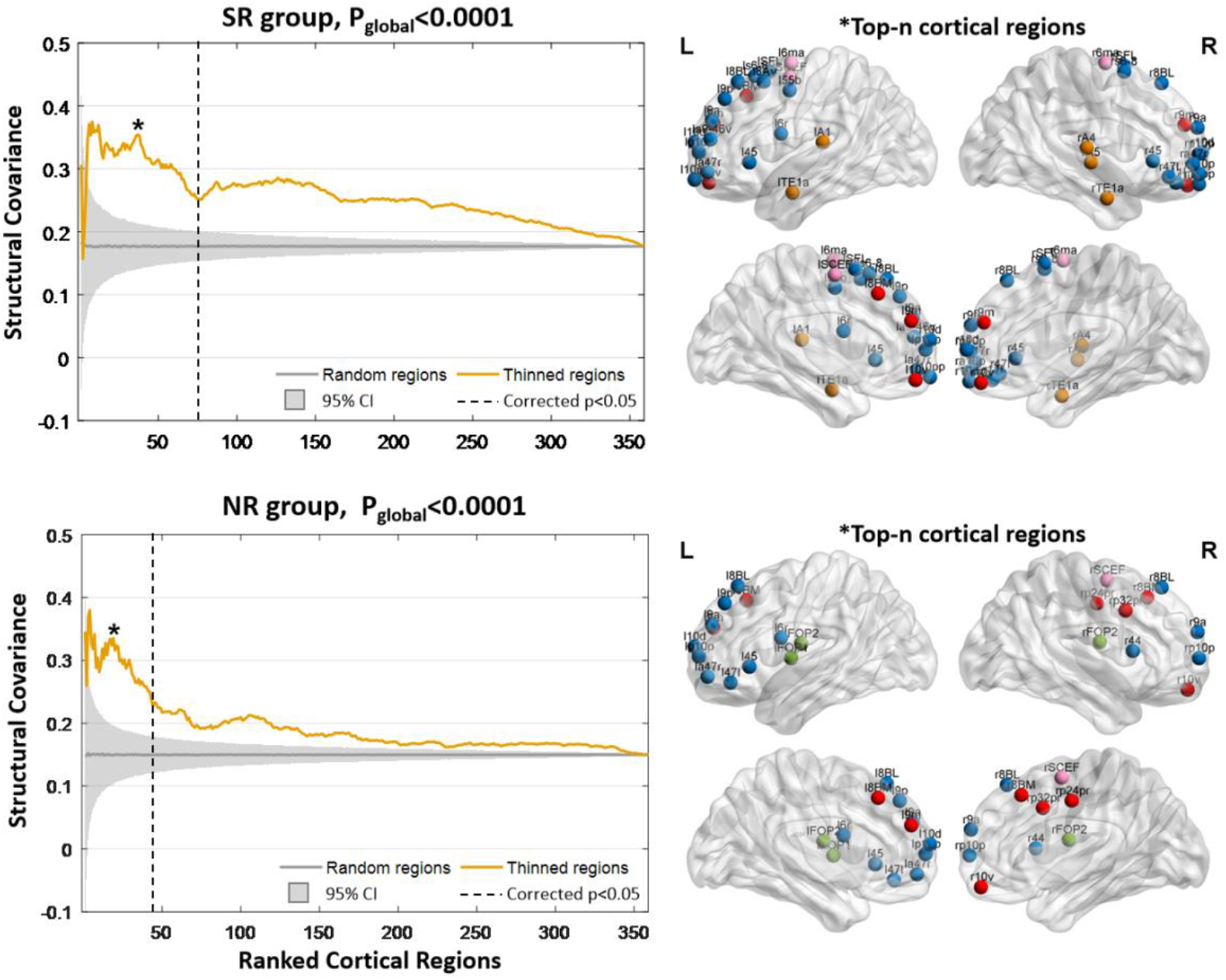
Structural covariance among the ranked regions with cortical thickness reductions in the SR and NR groups. The horizontal axis shows the cortical regions that ranked from high to low according to the severity of cortical thickness reductions using Cohen’d effect size. The dashed vertical line denotes the boundary between regions with and without significantly reduced cortical thickness after the antipsychotic medications (FWE corrected P<0.05). The vertical axis measures the structural covariance, which is computed as the Pearson’s correlation coefficient between two paired regions across the population of each group. The yellow line represents the mean structural covariance among the top-n ranked regions, i.e., SC_top-n_. The grey line indicates the mean structural covariance among the 5,000 randomly chosen n regions (i.e., SC_rand-n_). The shaded areas show the 95% confidence intervals across the permutated data, i.e., the 5,000 SC_rand-n_ values. The null hypothesis of equality in the area under curve (AUC) between the SC_top-n_ and SC_rand-n_ is rejected by the permutation test (P_global_<0.0001). Values of n marked with a star represent the strongest structural covariance (SC_top-n_) compared to SC_rand-n_ of the randomly chosen regions, which is determined by the SC_top-n_ z score (i.e., subtracting the average of SC_rand-n_ values and then dividing it by the standard deviation of SC_rand-n_ values). The brain node maps on the right side indicate the spatial distributions of top-n regions at the max SC_top-n_ z score. Nodes are colored according to cortex classifications (described in the Figure 2).

### SC in top-n regions with cortical thickness reduction

Figure 3 illustrates the mean SC among the top-n regions with the greatest extent of cortical thickness reduction. In the figure, the majority of SC_top-n_ values (the yellow line) exceed the shaded area (i.e., 95% confidence intervals of the average SC_rand-n_ values) (global P<0.0001), indicating that the SC between pairs of top-n regions was stronger than randomly chosen regions for both patient groups. In the NR group, the first 43 ranked regions showed decreased cortical thickness after treatment; when n=23, the top-n region presented the strongest SC, and the AUC at top-39 was 69.36. In the SR group, the first 77 ranked regions exhibited reduced cortical thickness at the follow-up. Of these regions, the top-n region was interconnected the most strongly when n was 39; the AUC at top-39 was 91.67. The majority of these top-n regions were located within the frontal and temporal regions and anterior cingulate cortex, especially for the right prefrontal and temporal regions, which were only observed in the SR group (Figure 3). Thus, SC across varying severity of cortical thickness reductions induced by antipsychotic medication was found to be increased in patients who responded to treatment compared with those who did not respond. Finally, the SC analysis in top n regions was also calculated in the HC group (Figure S3), which indicated that these regions with thickness changes following APM were strongly connected in the HC.

### Reproducibility and ancillary analysis

The ancillary analysis indicated that the SC results were repeatable (Figure S4). The group differences in baseline symptoms did not influence the SCN (Figure S5). The global efficiency difference between the SR and NR was not driven by the correlation strength (Figure S6). The between-group differences in top-n region SC did not result from potential geometric effects or hemispheric disproportion (Figure S7 and S8). Details are provided in the Supplementary Materials.

## Discussion

This study comprehensively investigated brain morphometry changes and then used a longitudinal study design to explore the short-term effects of atypical antipsychotics in medication-naive FES patients. First, regions with thinner cortical thickness were observed in the frontal, parietal, temporal and occipital lobes as well as the posterior cingulate cortex even in the early stage of the disease. Second, the effects of atypical antipsychotics were not constrained within a single brain region but exhibited a topographic distribution (i.e., network-level) across specific regions with cortical thickness reductions. The network-level assumption was demonstrated by the subsequent SCN approach, which identified stronger cortico-cortical connectivity and higher network integration in patients who responded to APM than among nonresponders. Third, the analysis of SC among top-n regions can provide further evidence supporting the link between stronger connectivity and thinner cortical thickness. Collectively, these findings support a potential network-level regulation mechanism of antipsychotics on brain structure, suggesting that the reconfiguration of morphological architecture induced by APM plays potential compensatory functions for abnormalities in the cerebral cortex.

### Thinner Cortical Thickness Are Found Even in the Early Stage of Medication-naive Patients with FES

Although thinner cortical thickness is a well-established feature in SZ, the degree of thinning has been demonstrated to vary across different illness stages ^4^. For example, FES patients show subtle cortical thinning mainly in the frontal and temporal cortices ^24^. In chronic SZ, pronounced reductions spread to parietal and occipital lobes ^25^. Individuals with treatment-resistant SZ exhibit more widespread cortical thickness thinning than patients who respond to treatment ^26^. Consistent with these studies ^8^, thinner cortical thickness was observed in the frontal, parietal, temporal and occipital lobes as well as posterior cingulate cortex in medication-naive FES patients. Interestingly, compared with healthy subjects, individuals who respond to treatment showed more widespread cortical thickness reductions at baseline than nonresponders, suggesting the potential association between pathological neuroanatomical changes and response to antipsychotic treatment ^27^. Supporting this result, relationships were found between volumes in frontal and medial temporal structures and treatment outcome ^28^. These findings support the existence of observable changes in morphometry of the cerebral cortex even in the SZ early stage.

### Cortical Thickness Reduction and Improved Structural Network Integrity Are the Short-term Effects of Atypical Antipsychotics

Accumulating evidence has demonstrated that APM could modulate brain morphology. More loss in gray matter volume has been reported in APM patients regardless of different analytic methods ^11^. Similarly, animal studies provide evidence of gray matter volume loss in macaque monkeys ^29^ and rodent exposure to antipsychotics ^30^. The loss in volume can be explained by fewer glial cells and higher densities of neurons and has been linked with neuroinflammatory models ^31, 32^. Conversely, some studies reported that APM increased gray matter volume in the prefrontal and occipital cortex in SZ ^33^. The inconsistency may be related to the illness stage, symptom severity, antipsychotic type, term of treatment, definition of response and analytic methods. Consistent with previous studies ^34^, this study observed widespread cortical thickness reductions regardless of treatment response. It is understandable when we assume that progressive neuroanatomical alterations are partly associated with the disease itself ^35^. Nonetheless, we found that some regions without baseline impairment compared with healthy controls exhibited cortical thickness reductions following APM only in the response group. This suggests that there are still morphological changes in specific areas that are due to antipsychotics effects independent of pathological progression.

In addition, the effects of antipsychotics on brain structure have been further demonstrated to occur not independently within constrained cortical locations but rather exhibit a topographically coherent pattern. This topographical pattern has been frequently linked with structural changes across development from childhood to early adulthood and continues across the lifespan ^36^. Abnormalities in structural covary patterns have been indicated to be involved in SZ pathophysiological processes and are associated with cortical gene expression in therapeutic targets and early brain development ^37^. Using a graph theoretical approach on a SCN, Palaniyappan and colleagues measured the structurally coherent pattern and found reduced global efficiency in SZ, indicating weaker structural network integrity ^5^. Hence, improved structural network integrity and more strongly cortico-cortical connectivity in responders highlight the potentially positive effects of antipsychotics on brain morphometry by means of connectomics, rather than traditionally interpreted as a detrimental effect if cortical thickness reduction was explored in isolation.

### Evidence supports the link between stronger connectivity and thinner cortical thickness

This study observed increased cortico-cortical connectivity between regions with thinner cortical thickness in FES patients, suggesting a link between connectivity and cortical morphological alterations. Supporting this finding, recent studies reported that cortical connectivity could shape and constrain the spatial distribution of cortical pathology in SZ ^4, 38^. A possible explanation is that local pathological processes, including abnormal neuronal signaling and neurotransmitter release, may propagate to distant regions by cortico-cortical connectivity ^4^. In patients with FES, abnormalities in morphology are related to disruptions in normal maturation and plasticity of higher order regions, such as the prefrontal, parietal and temporal cortices ^39^, which have also presented pathological decreases in these regions in pre-APM patients. Notably, stronger SC between thinned regions was found in the responders after taking APM. In addition, the covariance network of nonresponders was found to lack key nodes of the right prefrontal and temporal regions compared with that of responders, indicating better reconfiguration of morphological architecture in APM responders. These findings suggest that structural abnormalities of the cerebral cortex in FES are balanced by the reconfiguration of morphological architecture and increasing interregional covariance, possibly reflecting a compensation process ^5^ when symptoms are controlled by antipsychotics.

### Limitations

Several limitations must be considered. First, although the 12-week follow-up investigated the short-term effect of APM on brain structure, the long-term persistence of antipsychotics is still a question. Meanwhile, although we assume that brain structural changes over 12 weeks are subtle in healthy adults, normal neurodevelopment or neurodegeneration progression cannot be neglected from the presented results due to the lack of follow-up in healthy controls. Second, the definition of treatment response is based on a cutoff of 50% PANSS reductions. Other cutoff points (i.e., <30%, 30-50% and 50-100%) are also used. Third, the choice of antipsychotic medications is clinically led. The current study could not separate the specific effects of different antipsychotic types. In addition, the adipogenic effect of antipsychotics is a potential bias, as weight gain is evident in FES patients after medication treatment. It is also difficult to rule out the possible effects of variations in brain microstructure (i.e., myelination, iron, and water content) on cortical thickness reductions ^40^. Due to the lack of cognitive assessments, the relationship between the structural network and cognitive deficits in SZ was not examined in this study.

## Conclusions

The APM induced more cortical thickness reductions, even in the frontotemporal regions without baseline reductions, thereby reflecting the specific effects of antipsychotics independent of pathological progression. Furthermore, antipsychotic-induced changes in brain structures have been demonstrated to occur not independently within constrained cortical locations but rather exhibit a network-level SC pattern. The relationship between increased SC and cortical thickness reductions suggests that abnormalities in cortical morphology may be compensated by increasing interregional covariance when symptoms are controlled by antipsychotics. Patients who responded to medications showed stronger cortico-cortical connectivity and higher network integration after APM, thus highlighting the potentially positive effects of antipsychotics on the reconfiguration of brain morphological architecture by means of connectomics.

## Data Availability

The data supporting the findings of this study are available from the corresponding author upon reasonable request.

## Acknowledgements

This work was partly supported by the grant from the National Key R&D Program of China (No. 2018YFA0701400), grants from the National Natural Science Foundation of China (No. grant number: 61933003, 81771822, 81861128001, and 81771925), the CAMS Innovation Fund for Medical Sciences (CIFMS) (No. 2019-I2M-5-039) and the Project of Science and Technology Department of Sichuan Province (No. 2019YJ0179). We thank the American Journal Experts Editing (https://www.aje.com/) for editing the English of the manuscript.

## Disclosures

There is no conflict of interest.

## Notes

### Competing Interest Statement

The authors have declared no competing interest.

### Clinical Trial

This study is an observational study, not a randomized clinical trial

### Author Declarations

The study was approved by the Institutional Review Board of Shanghai Mental Health Center. All subjects/patients signed written informed consent forms.

