## Supplementary Materials for "Antipsychotics effects on network-level reconfiguration of cortical morphometry in first-episode schizophrenia"

### *Participants information*

A total of 127 patients with medication-naive FES (SZ group) who were diagnosed using the Diagnostic and Statistical Manual of Mental Disorders, 4th Edition (DSM-IV) and 133 healthy controls (HC group) matched for age, gender, education and handedness were recruited from Shanghai Mental Health Center. The exclusion criteria were as follows: (1) brain trauma, substance-related disorders, major medical or neurologic disorders; (2) other mental disorders meeting DSM-IV criteria; (3) drug or alcohol abuse; (4) pregnancy, breastfeeding or other unstable clinical state including aggressive behavior; (5) history of electroconvulsive therapy or transcranial magnetic stimulation within six months; and (6) other contraindications to MRI scanning. To eliminate potential familial effects, healthy subjects whose first- or second-degree relatives had a history of mental disorders were also excluded. MRI was scanned at baseline for all subjects and at follow-up after 12 weeks of APM for only patients. Following the baseline scanning, patients received second-generation antipsychotics. The daily dosage was converted to chlorpromazine equivalents and described in the following Supplementary Table 1. The severity of symptoms was evaluated at baseline and then at the 12-week follow-up using the Positive and Negative Syndrome Scale (PANSS) by the same psychiatrist. Symptom relief was measured using the reduction in PANSS total scores defined by the following formula [1]: $\triangle PANSS=(PANSS_{t1}-PANSS_{t2})\times 100\%/(PANSS_{t1}-30)$. Patients were grouped as responders (SR group, n=75) and nonresponders (NR group, n=52) by the criterion of less than 50% PANSS reduction for nonresponse [2]. The study was approved by the Institutional Review Board of Shanghai Mental Health Center. All subjects/patients signed written informed consent forms.

### *Medications for patients*

All the patients received atypical antipsychotics, ninety three patients received monotherapy: olanzapine (n=32), risperidone (n=18), aripiprazole (n=17), amisulpride (n=16), paliperidone (n=8), quetiapine (n=2) and ziprasidone (n=1).Thirty-four patients received combined therapy: aripiprazole and olanzapine (n=9), aripiprazole and risperidone (n=5), amisulpride and olanzapine (n=4), risperidone and olanzapine (n=4), risperidone and quetiapine (n=3), aripiprazole and paliperidone (n=2), quetiapine and paliperidone (n=2), aripiprazole and quetiapine (n=1), ziprasidone and olanzapine (n=1), and ziprasidone and aripiprazole (n=1), amisulpride and paliperidone (n=1), The average dose in chlorpromazine equivalence (CPZ eq) was 402.78±188.96 mg/day [3]. Information on antipsychotic medication usage for each patient are provided in the following Table 1.

**Supplementary Table 1. Information on antipsychotic medication usage for each patient.**

| Subject | Group* | Drug 1 | Average Dose | Drug 2 | Average Dose | CPZ equivalent units |
|---|---|---|---|---|---|---|
| sub1 | 1 | Risperidone | 4 | Aripiprazole | 10 | 440 |
| sub2 | 1 | Olanzapine | 30 | | | 900 |
| sub3 | 0 | Risperidone | 4 | | | 240 |
| sub4 | 0 | Olanzapine | 15 | Aripiprazole | 10 | 650 |
| sub5 | 0 | Aripiprazole | 15 | Risperidone | 3 | 480 |
| sub6 | 1 | Amisulpride | 400 | | | 300 |
| sub7 | 1 | Olanzapine | 5 | | | 150 |
| sub8 | 1 | Olanzapine | 20 | | | 600 |
| sub9 | 1 | Risperidone | 5 | | | 300 |
| sub10 | 0 | Olanzapine | 20 | | | 600 |
| sub11 | 0 | Aripiprazole | 5 | | | 100 |
| sub12 | 0 | Amisulpride | 400 | | | 300 |
| sub13 | 0 | Olanzapine | 10 | | | 300 |
| sub14 | 1 | Aripiprazole | 15 | | | 300 |
| sub15 | 0 | Aripiprazole | 20 | | | 400 |
| sub16 | 1 | Risperidone | 3 | | | 180 |
| sub17 | 1 | Olanzapine | 15 | | | 450 |
| sub18 | 1 | Aripiprazole | 5 | | | 100 |
| sub19 | 1 | Olanzapine | 5 | | | 150 |
| sub20 | 0 | Amisulpride | 200 | | | 150 |
| sub21 | 1 | Risperidone | 3 | | | 180 |
| sub22 | 1 | Risperidone | 4 | | | 240 |
| sub23 | 1 | Risperidone | 4 | | | 240 |
| sub24 | 0 | Olanzapine | 10 | | | 300 |
| sub25 | 0 | Aripiprazole | 20 | | | 400 |
| sub26 | 0 | Paliperidone palmitate injection | 100 | | | 428.57 |
| sub27 | 0 | Olanzapine | 10 | | | 300 |
| sub28 | 1 | Aripiprazole | 20 | | | 400 |
| sub29 | 0 | Aripiprazole | 15 | | | 300 |
| sub30 | 0 | Olanzapine | 15 | | | 450.45 |
| sub31 | 0 | Quetiapine | 400 | | | 300 |
| sub32 | 0 | Risperidone | 8 | | | 480 |

| | | | | | | |
|---|---|---|---|---|---|---|
| sub33 | 0 | Aripiprazole | 5 | | | 100 |
| sub34 | 0 | Amisulpride | 600 | | | 450 |
| sub35 | 0 | Olanzapine | 10 | | | 300 |
| sub36 | 1 | Olanzapine | 15 | | | 450 |
| sub37 | 0 | Amisulpride | 400 | | | 300 |
| sub38 | 0 | Risperidone | 3 | Aripiprazole | 10 | 380 |
| sub39 | 0 | Risperidone | 4 | | | 240 |
| sub40 | 0 | Olanzapine | 20 | Amisulpride | 100 | 675 |
| sub41 | 1 | Amisulpride | 200 | | | 150 |
| sub42 | 0 | Olanzapine | 5 | | | 150 |
| sub43 | 1 | Risperidone | 3 | | | 180 |
| sub44 | 0 | Risperidone | 6 | Aripiprazole | 3.33 | 427 |
| sub45 | 0 | Amisulpride | 200 | | | 150 |
| sub46 | 0 | Aripiprazole | 10 | Quetiapine | 800 | 800 |
| sub47 | 0 | Amisulpride | 600 | | | 450 |
| sub48 | 0 | Olanzapine | 10 | | | 300 |
| sub49 | 0 | Olanzapine | 20 | | | 600 |
| sub50 | 1 | Olanzapine | 15 | | | 450 |
| sub51 | 1 | Amisulpride | 600 | | | 450 |
| sub52 | 1 | Olanzapine | 15 | | | 450 |
| sub53 | 1 | Amisulpride | 500 | | | 375 |
| sub54 | 1 | Olanzapine | 10 | Amisulpride | 200 | 450 |
| sub55 | 1 | Risperidone | 5.5 | | | 330 |
| sub56 | 0 | Olanzapine | 20 | Aripiprazole | 20 | 1000 |
| sub57 | 0 | Quetiapine | 400 | Risperidone | 2 | 420 |
| sub58 | 1 | Ziprasidone | 60 | Aripiprazole | 10 | 425 |
| sub59 | 1 | Risperidone | 3 | | | 180 |
| sub60 | 1 | Amisulpride | 200 | | | 150 |
| sub61 | 0 | Olanzapine | 15 | | | 450 |
| sub62 | 1 | Olanzapine | 5 | Aripiprazole | 10 | 350 |
| sub63 | 1 | Olanzapine | 20 | | | 600 |
| sub64 | 1 | Amisulpride | 200 | | | 150 |
| sub65 | 1 | Aripiprazole | 20 | | | 400 |
| sub66 | 1 | Olanzapine | 10 | Aripiprazole | 20 | 700 |
| sub67 | 0 | Olanzapine | 5 | | | 150 |
| sub68 | 1 | Olanzapine | 5 | | | 150 |
| sub69 | 1 | Amisulpride | 400 | Olanzapine | 20 | 900 |
| sub70 | 0 | Olanzapine | 10 | | | 300 |
| sub71 | 1 | Aripiprazole | 5 | Olanzapine | 10 | 400 |
| sub72 | 0 | Olanzapine | 10 | | 300 | 300 |

| | | | | | | |
|---|---|---|---|---|---|---|
| sub73 | 1 | Aripiprazole | 5 | | 100 | 100 |
| sub74 | 1 | Olanzapine | 10 | | | 300 |
| sub75 | 1 | Olanzapine | 10 | | | 300 |
| sub76 | 0 | Amisulpride | 400 | | | 300 |
| sub77 | 1 | Aripiprazole | 10 | Olanzapine | 3.33 | 300 |
| sub78 | 1 | Amisulpride | 200 | Olanzapine | 5 | 300 |
| sub79 | 1 | Olanzapine | 20 | Risperidone | 6 | 960 |
| sub80 | 0 | Risperidone | 5 | | | 300 |
| sub81 | 1 | Amisulpride | 400 | | | 300 |
| sub82 | 1 | Aripiprazole | 10 | | | 200 |
| sub83 | 1 | Olanzapine | 15 | Aripiprazole | 20 | 850 |
| sub84 | 0 | Paliperidone | 9 | Amisulpride | 133.33 | 550 |
| sub85 | 0 | Amisulpride | 600 | | | 450 |
| sub86 | 0 | Aripiprazole | 20 | | | 400 |
| sub87 | 1 | Ziprasidone | 100 | | | 375 |
| sub88 | 1 | Olanzapine | 10 | Risperidone | 2 | 420 |
| sub89 | 1 | Paliperidone | 12 | | | 600 |
| sub90 | 0 | Olanzapine | 20 | | | 600 |
| sub91 | 1 | Amisulpride | 800 | | | 600 |
| sub92 | 0 | Aripiprazole | 15 | | | 300 |
| sub93 | 1 | Aripiprazole | 25 | | | 500 |
| sub94 | 1 | Risperidone | 8 | | | 480 |
| sub95 | 1 | Olanzapine | 20 | | | 600 |
| sub96 | 1 | Aripiprazole | 10 | | | 200 |
| sub97 | 1 | Olanzapine | 10 | | | 300 |
| sub98 | 1 | Aripiprazole | 20 | | | 400 |
| sub99 | 0 | Risperidone | 6 | | | 360 |
| sub100 | 1 | Olanzapine | 20 | | | 600 |
| sub101 | 1 | Risperidone | 4 | Aripiprazole | 10 | 440 |
| sub102 | 1 | Paliperidone | 6 | | | 300 |
| sub103 | 1 | Risperidone | 2.5 | Quetiapine | 800 | 750 |
| sub104 | 1 | Olanzapine | 15 | Aripiprazole | 10 | 650 |
| sub105 | 0 | Aripiprazole | 20 | | | 400 |
| sub106 | 1 | Olanzapine | 12.5 | Ziprasidone | 20 | 450 |
| sub107 | 1 | Paliperidone | 4 | Aripiprazole | 2.5 | 450 |
| sub108 | 0 | Quetiapine | 100 | Paliperidone | 4.5 | 300 |
| sub109 | 1 | Olanzapine | 7.5 | Quetiapine | 200 | 375 |
| sub110 | 1 | Risperidone | 2 | Olanzapine | 20 | 720 |
| sub111 | 1 | Paliperidone | 9 | | | 450 |
| sub112 | 1 | Paliperidone | 100 | | | 428.57 |

| | | | | | | |
|---|---|---|---|---|---|---|
| | | palmitate injection | | | | |
| sub113 | 1 | Risperidone | 4 | | | 240 |
| sub114 | 1 | Risperidone | 3 | | | 180 |
| sub115 | 1 | Olanzapine | 15 | Risperidone | 1 | 510 |
| sub116 | 1 | Paliperidone | 6 | Aripiprazole | 5 | 400 |
| sub117 | 1 | Olanzapine | 5 | Aripiprazole | 20 | 550 |
| sub118 | 0 | Risperidone | 6 | | | 360 |
| sub119 | 0 | Paliperidone | 9 | Quetiapine | 300 | 675 |
| sub120 | 0 | Paliperidone | 9 | | | 450 |
| sub121 | 1 | Olanzapine | 10 | | | 300 |
| sub122 | 1 | Olanzapine | 20 | | | 600 |
| sub123 | 1 | Risperidone | 4.5 | | | 270 |
| sub124 | 1 | Paliperidone palmitate injection | 150 | | | 642.86 |
| sub125 | 1 | Paliperidone | 9 | | | 450 |
| sub126 | 0 | Quetiapine | 700 | | | 525 |
| sub127 | 0 | Olanzapine | 20 | | | 600 |

Note: *Group, 1=Responders; 0=Nonresponders.

### *Image Data Acquisition*

A 3-Tesla MRI scanner (Siemens MR B17) was used to acquire data in the Shanghai Mental Health Centre. High-spatial-resolution T1-weighted images were collected by a magnetization-prepared rapid acquisition gradient echo (MPRAGE) sequence. The main parameters included repetition time /echo time, 2530/2.56 msec; flip angle, 7°; field of view, 256 × 256 $mm^2$; matrix size, 256 × 256; section thickness, 1 mm (no gap); voxel size, 1×1×1$mm^3$.

### *Permutation-based Statistical Testing*

To correct for multiple comparisons and to minimize the bias of data distribution, a permutation-based method was employed [4]. The permutation-based statistical testing permits the inference of the probability of the observed statistic, such as a t-value from a distribution of the same statistic estimated from massive instances of the same samples with their group identities permuted [5]. We can expand the traditional method by applying a $t_{max}$ principle to adjust the estimated P-value of observations for multiple comparisons by controlling the FWE rate [6]. In this case, a permutation testing procedure begins the same way as the traditional approach, i.e., by computing a statistic of *t-value* for the observed data in the sample A and B, *t-observed*. Next, a permutation step is conducted by randomly swapping labels of observations and a new *t*-score is recomputed for the exchanged two samples under this randomly rearrangement. The permutation process is repeated 100,000 times and thus generated a distribution of the possible *t-values* under the null hypothesis that observations are exchangeable. According to the location of *t-observed* in the distribution of the possible *t-values*, the P-value is estimated to how probable such observations would be if the null hypothesis is true. This method can correct for multiple comparisons because the distribution of the most extreme statistics automatically adjusts to reflect the increased chance of false discoveries due to an increased number of comparisons [4].

### *Associations between Cortical Thickness and treatment outcomes*

In the main text, regression analysis found a significant relationship between the cortical thickness change in the left area-45 of inferior frontal cortex and PANSS total reduction (T=2.37, P<0.05) in the SR group (Figure S1). To further investigate whether the association was affected by confounding factors, partial correlation analysis was used to further assess the correlation between the cortical thickness change in the left area-45 of inferior frontal cortex and PANSS total reduction, when controlling for the effects of the demographic (gender, age, education and TIV), clinical information (CPZ and illness duration) or symptom (baseline PANSS total scores), separately. Partial correlation analysis demonstrated that the correlation remained significant after controlling the demographic (R=0.275, P=0.02), clinical information (R=0.274, P=0.019) or symptom (R=0.268, P=0.021).

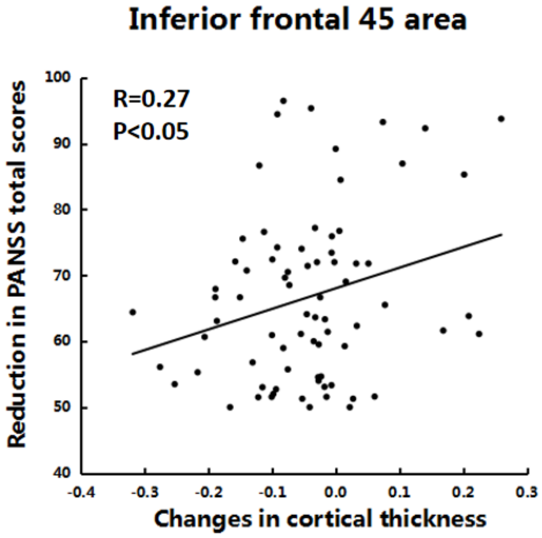


**Figure S1.** The scatter plot shows a significant association between the cortical thickness change in the left area-45 of inferior frontal cortex and PANSS total reduction (R=0.27, P<0.05) in the SR group.

### *Structural covariance network*

To further investigate the network integration of the structural covariance network, global efficiency was used for the measurement of network topology. The global efficiency of a network is denoted by the averaged inverse shortest path length and has been regarded as a superior measure of integration. The null hypothesis of equality in the global efficiency of the structural covariance network between the SR and NR groups was tested using a permutation test. In brief, we calculated the real difference value in global efficiency between the two groups. The group labels were randomly assigned across all patients. The structural covariance network was reconstructed in each randomized group. We recalculated the difference in the global efficiency of the two random structural covariance networks. This randomization procedure was repeated 10,000 times and thus yielded a distribution of observations under the null hypothesis. Based on the location of the real difference value within the distribution of the null hypothesis, a p-value was assigned to the real difference.

In the structural covariance network of SR group, majority of these connections (63.04%) were associated with frontal and parietal cortices; however, the frontal and parietal connections account for 45.78% of all connections in the NR group, as shown in Figure S2.

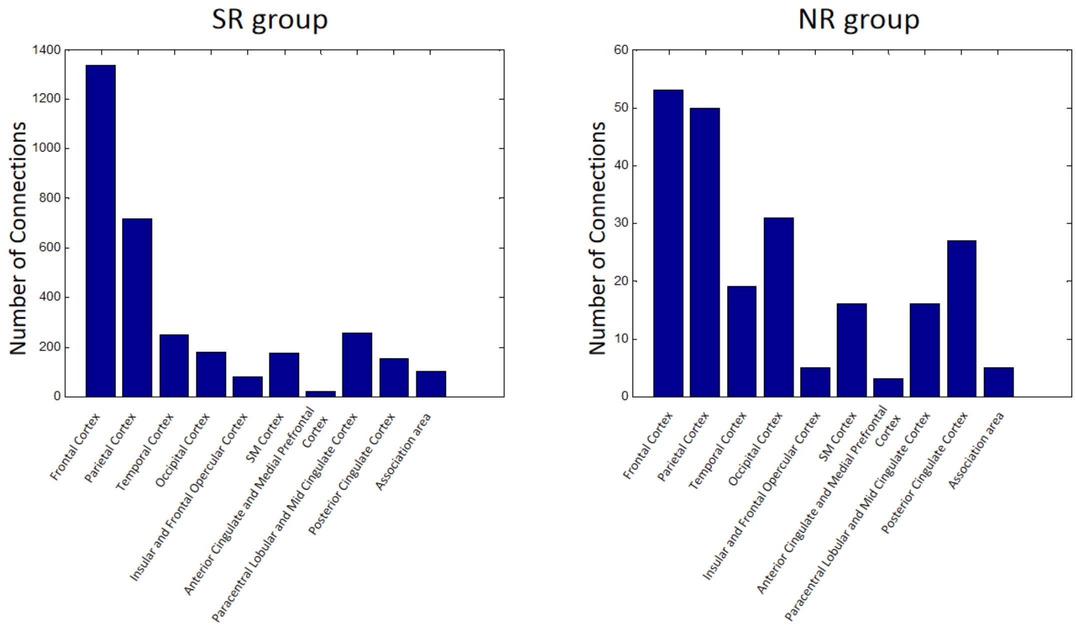


**Figure S2.** The number of connections with different brain areas in the structural covariance networks of the SR and NR groups.

#### *Structural covariance in top-n regions*

By sorting the regions with thickness changes after medications from high to low, the structural covariance within the top n regions reflects the relatively similar rate of thickness changes over time in these regions. To further examine whether these regions are connected in healthy controls. We computed the structural covariance of top n region in the HC group. In these regions with cortical thickness changes after treatment, there are strongly structural covariance in the HC group (P<0.05) (Figure S3). This indicated these regions are connected in the HC.

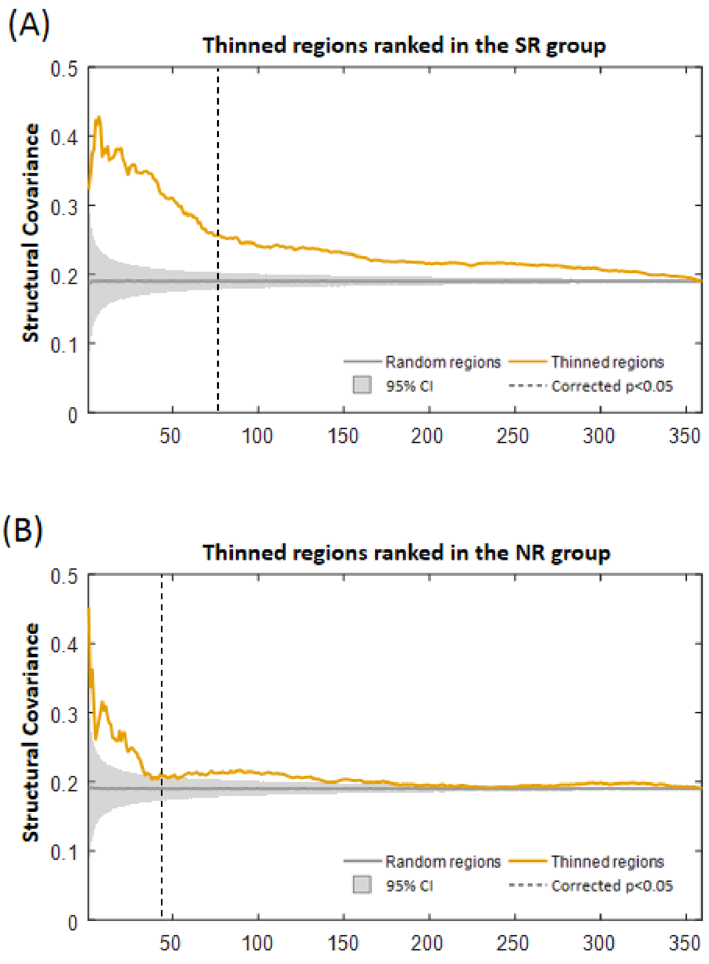


**Figure S3.** Structural covariance in the HC group. (A) The regions were ranked according to the thickness changes in the SR group. (B) The regions were ranked according to the thickness changes in the NR group.

### *Reproducibility and ancillary analysis*

In ancillary analysis 1, to evaluate the reproducibility of the structural covariance network, subjects in each patient cohort were randomly divided into two subsamples. The structural covariance network was recomputed in each random subsample and compared with each other. In ancillary analysis 2, the structural covariance network was repeated with the baseline PANSS general score as a covariate. This step was taken to eliminate the potential differences in baseline symptom scales between the two patient groups. In ancillary analysis 3, the average strength of the structural covariance network was compared between groups. We also analyzed the global efficiency when controlling for the correlation strength differences between groups. In ancillary analysis 4, we investigate the potential impact of the geometric effect and hemispheric disproportion on the structural covariance of top-n regions between two patient groups.

### *Ancillary analysis 1*

To evaluate the reproducibility of structural covariance network, subjects in each patient cohort were randomly divided into two subsamples. In each random subsample, structural covariance network was re-computed using the same method mentioned in the body text. Pearson's correlation coefficient was used for the quantification of similarities between the structural covariance networks of two subsamples. The above similarity analysis was repeated 100 times. The averaged correlation coefficient was 0.385 ($P<0.00001$) in the SR group (Figure S4). The averaged correlation coefficient was 0.226 ($P<0.00001$) in the NR group (Figure S4). To further evaluate the potential bias in intercohort sample size, we compare the similarity between the full sample and each random subsample. The averaged correlation coefficient was 0.806 ($P<0.00001$) in the SR group and 0.780 ($P<0.00001$) in the NR group. In summary, structural covariance network showed satisfactory consistency in the SR group ($R=0.385$, $P<0.00001$) and significant consistency ($R=0.226$, $P<0.00001$) in the NR group. In addition, intercohort sample size did not introduce bias on these structural covariance network.

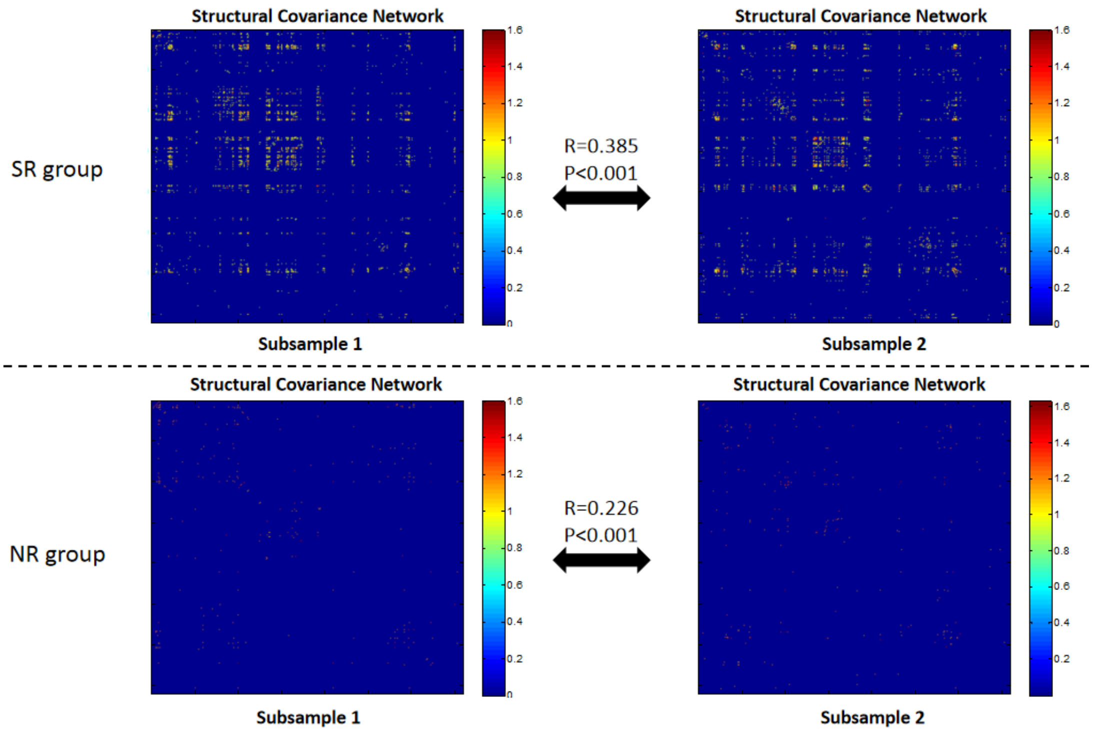


**Figure S4.** Reproducibility evaluation of structural covariance network indicated satisfactory consistency in the SR group (R=0.385, P<0.001) and significant consistency (R=0.226, P<0.001) in the NR group.

### *Ancillary analysis 2*

As higher scores in the baseline PANSS general scale were observed in the SR group, structural covariance network was re-computed with the addition of the baseline PANSS general score as a covariate to eliminate the potential bias in baseline symptom. Pearson's correlation coefficient was used to quantitatively assess the consistency between the structural covariance networks including or not including the baseline PANSS general score as a covariate. We found that the structural covariance network remained highly similar pattern in either including the baseline PANSS general score as a covariate or not (see Figure S5). The high similarity was observed in both two patient groups (SR group, R=0.895, P<0.001; NR group, R=0.871, P<0.001), suggesting that the group differences in baseline symptom did not introduce bias on the structural covariance network.

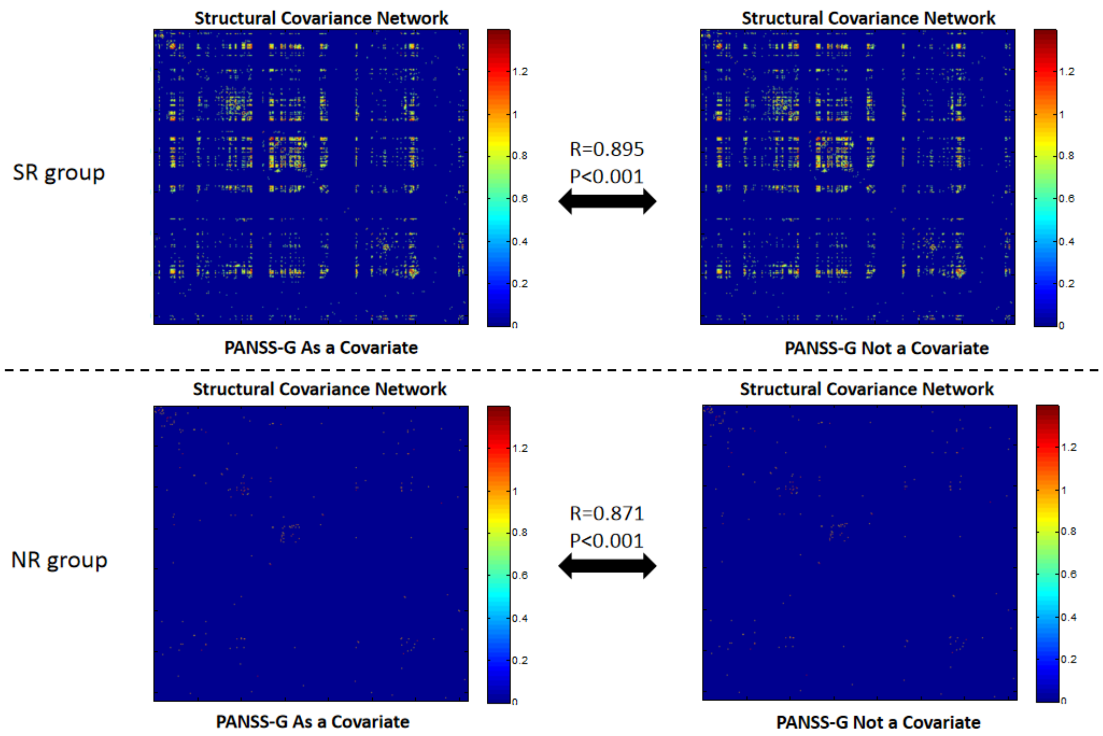


**Figure S5.** Structural covariance network remained highly similar pattern in either including the baseline PANSS general score (PANSS-G) as a covariate or not.

### *Ancillary analysis 3*

We compared the average correlation strength of the structural covariance network between the SR and NR groups using the permutation test. We found that there was a significant difference between the two groups (P<0.001) (Figure S6 A).

To further identify whether the group difference in the global efficiency was driven by the correlation strength difference, we re-analyzed the global efficiency when controlling for the correlation strength differences between groups. The normalized global efficiency was defined as follows:

$$\text{Normalized global efficiency} = \frac{\text{global efficiency}}{\text{averaged correlation strength}}$$

We compared the normliazed global efficiency between the SR and NR using the permutation test. We found that the normalized global efficiency in the SR group was significantly higher than in the NR group (Figure S6 B), which is consistent with the results in the main text. This indicated that the global efficiency difference between the SR and NR was not driven by the correlation strength.

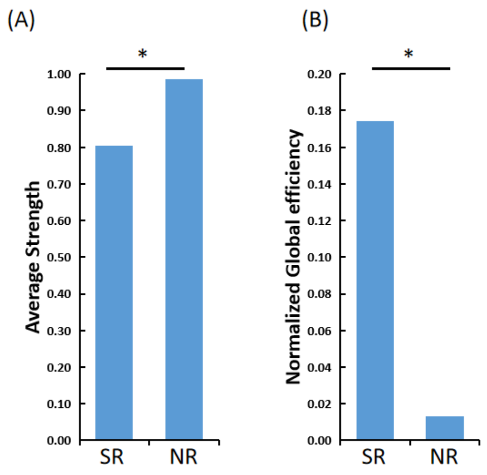


**Figure S6.** The differences in the averaged strength (A) and normalized global efficiency (B) between the SR and NR groups by the permutation test.

### *Ancillary analysis 4*

To investigate the potential impact of the geometric effect on the structural covariance of top-n regions with cortical thickness reduction, we computed the mean Euclidean distance between pairs of top-n regions with cortical thickness reductions. This was plotted as a function of n from 2 to 360 in the SR and NR group, respectively (see Figure S7). In addition, the number of top-n regions within each cerebral hemisphere was also plotted as a function of n to illustrate the potential impact of the hemispheric disproportion on the structural covariance of top-n regions (see Figure S8). We observed that the mean Euclidean distance across top-n regions exhibited similar pattern in the two patient groups (R=0.965, P<0.001). Also, the number of top-n regions within each cerebral hemisphere showed similar pattern in the two patient groups (R=0.997, P<0.001). These findings confirmed that the between-group differences in top-n regions structural covariance were not the simply results from potential geometric effect or hemispheric disproportion.

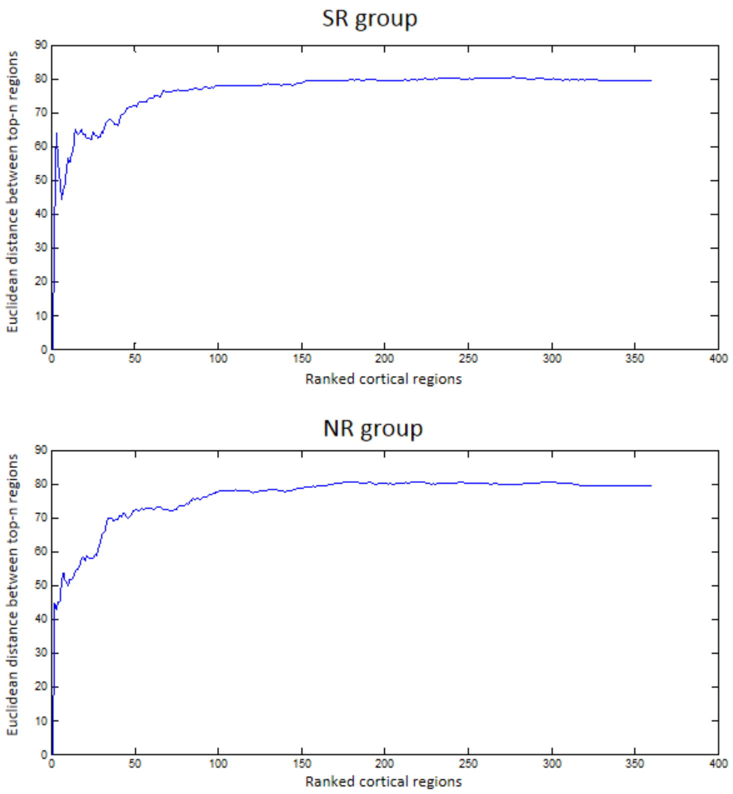


**Figure S7.** The mean Euclidean distance between pairs of top-n regions with cortical thickness reductions.

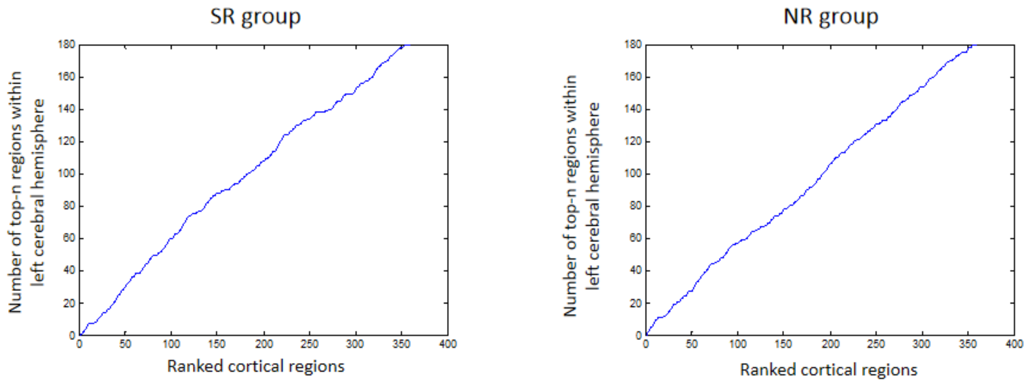


**Figure S8.** The number of top-n regions within each cerebral hemisphere.
